# Mode of Presentation and Outcomes of COVID-19 Cases in a Tertiary Hospital in Nigeria

**DOI:** 10.1101/2021.07.06.21260084

**Authors:** Yakubu Egigogo Raji, Bala Waziri, Sadiq Aliyu Hussaini, Ahmad Idris Ja’agi, Umar Isah Alhaji, Abdulmalik M. Aliyu, Abdullahi Muhammad, Adama Saidu Garba

## Abstract

Coronavirus disease 2019 (COVID-19) has spread across the globe with its consequent human and economic challenges. To achieve effective control of the pandemic, efforts need to be holistic and global. Understanding patients’ demographics and clinical characteristics will assist in the control of the infection. However, there is a paucity of studies on the clinical presentation of COVID-19 patients from Nigeria and indeed Africa. Thus, this retrospective case series evaluated the medical records of COVID-19 patients admitted in a tertiary hospital in Nigeria. Patients’ demographics, and other clinical variables were assessed and presented. Data of 14 patients with complete records were included in the study. Most of the patients (78.6%) were males and the mean age of the study participants is 63.5 years (SD; 11.5). The commonest presenting symptoms were fever (93%), cough (71.4%), and dyspnoea (57.1%). At presentation, 13 patients had coexisting diseases while 8 (57.0%) patients had moderate disease and the remaining 6 (43.0%) had severe cases. After management, 1 patient died, two were referred and 11 recovered and were discharged alive. Thus, this study has identified advanced age, male gender, and comorbidity as increased risk factors for hospitalisation. The patient survival outcome in this study was also good.

## Introduction

In December 2019 a cluster of life-threatening pneumonia cases emerged in the Wuhan city of China [1]. Afterward, in early January 2020, the genetic sequence of the agent causing the disease was identified to be a novel coronavirus [2]. Thus, the virus was temporarily named the 2019 novel coronavirus (2019-nCov) [2]. Later in February 2020, the World Health Organisation (WHO), named the disease caused by the 2019-nCoV the “Coronavirus disease 2019” (COVID-19) [3]. The International Committee on Taxonomy of Viruses (ICTV) subsequently named the virus as the ‘severe acute respiratory syndrome-related coronavirus-2 (SARS-CoV-2) [4].

Coronaviruses are enveloped viruses of approximately 80 to 200 nanometres in diameter. The viruses have a single-stranded RNA genome of 26 to 32 kilobases. Coronaviruses belong to the order, family, and subfamily of *Nidovirales, Coronaviridae*, and *Coronavirinae* respectively. The subfamily Coronavirinae has four genera: *Alphacoronavirus, Betacoronavirus, Gammacoronavirus*, and *Deltacoronavirus*. The viruses in the *Alphacoronavirus* and *Betacoronavirus* genera are primarily human coronaviruses (HCoVs). Previously known HCoVs as established human pathogens include HCoV-NL63, HCoV-229E, (Alphacoronaviruses) HCoV-HKU1, HCoV-OC43, Middle East respiratory syndrome coronavirus (MERS-CoV), and SARS-CoV (Betacoronaviruses) [5]. The first four of these viruses are known for causing mild upper respiratory disease. While the last two can lead to severe and lethal respiratory disease.

Also, SARS-CoV-2 the agent causing COVID-19 belongs to the Betacoronavirus genus and is the seventh known HCoV. Similarly, COVID-19 clinical presentation ranges from mild (or asymptomatic) illness to severe (or critical) disease and even death. Some of the common clinical manifestations are fever, cough, dyspnoea, myalgia, fatigue, sore throat, headache, anosmia, nausea, vomiting, and diarrhoea [6]. The severe form of the disease is often accompanied by complications. Such complications include acute respiratory distress syndrome (ARDS), acute liver injury, acute cardiac injury, and acute kidney injury amongst others.

Since the emergence of this disease, it has spread across the globe affecting virtually all countries in the world. Thus, causing global human development crises and economic challenges. As in many other countries, Nigeria was also affected by the COVID-19 pandemic. The first COVID-19 case recorded in Nigeria was a visiting Italian on the 27^th^ day of February 2020 [7]. Subsequently, the pandemic has affected all the 36 states of Nigeria and the federal capital [8]. As at the time of writing, the country has recorded over 160,000 cases with more than 2000 deaths [8]. However, despite this huge number of reported cases with likely different modes of presentation, there is still a paucity of literature on patients’ clinical characteristics in Nigeria. Thus, there is a need for more studies on the clinical characteristics of COVID-19 patients in Nigeria. Therefore, this study reports the characteristics and clinical presentation of COVID-19 patients in a tertiary hospital in Nigeria.

## Case Presentation

The cases reported in this study were of COVID-19 patients treated at Ibrahim Badamasi Babangida Specialist Hospital (IBBSH). IBBSH is a 100 bedded tertiary hospital located in Minna, the capital city of Niger state in North-central Nigeria. The hospital is comprised of departments of internal medicine, family medicine, general surgery, urology, orthopaedic surgery, radiology, and obstetrics, and gynaecology. As a tertiary hospital, IBBSH serves as the major referral centre in the state.

Cases were collected retrospectively from patients’ medical records. Patients included in this case series are those that presented between 30^th^ June 2020 to 28^th^ January 2021. Data collected from patient’s medical records include demographics, exposure history, clinical presentations, radiology, and laboratory investigations, clinical management, and outcome. Only patients with complete variables as outlined above were included in this report. Before the commencement of this study, ethical approval was obtained from the hospital ethics and review committee. To ensure confidentiality, non-authorised personnel had no access to patient medical records. Also, all patients’ data were retrieved by clinicians and anonymously recorded.

At the point of presentation to the hospital, all patients were screened for the identification of suspected COVID-19 cases. Screenings were conducted following the WHO interim guideline for the clinical management of COVID-19 [9]. Identified suspected patients were isolated from the general patients at the hospital isolation centre. Samples were then collected per the Nigeria Centre for Disease Control (NCDC) guideline [10]. Collected samples (oropharyngeal and nasopharyngeal swabs) were then transported to the National Reference Laboratory (NRL) Abuja. At the NRL, COVID-19 real-time reverse-transcription polymerase chain reaction (RT-PCR) test was conducted on each of the samples. All patients included in the study had positive SARS-CoV-2 RT-PCR test results in accordance with NCDC guidelines [10].

Included in this report, are a total of 14 patients that were admitted and treated for COVID-19 at IBBSH from 30^th^ June 2020 to 28^th^ January 2021. The 14 patients included were those with complete data of the variables of interest. More than a three-quarter (78.6%) of the patients were males. The mean age (in years) of the study participants is 63.5 (SD; 11.5).

All the 14 patients were treated as in-patients and 10/14 (71.4%) were admitted through the accident and emergency (A&E) unit of the hospital. While the remaining four patients were admitted through the out-patient unit. Out of the 10 patients admitted via the A&E unit, three (30%) were referred cases from private hospitals. At the referral hospitals, the three patients were either managed for malaria and/or chest infection. The median duration of onset of symptoms and admission at the facility was 4 days (Interquartile range [IQR]: 2.75-8.75). Patients presented with varied symptoms and physical examination findings. Details of the clinical presentations are given in table 1. The commonest presenting symptoms were fever (93%), cough (71.4%), and dyspnoea (57.1%). Six patients out of the 14 (43%), had a history of contact; one of the patients had contact with a confirmed case, another one had a history of travel and the remaining four were all referred from healthcare facilities where COVID-19 case(s) have been reported.

**Table 1:**
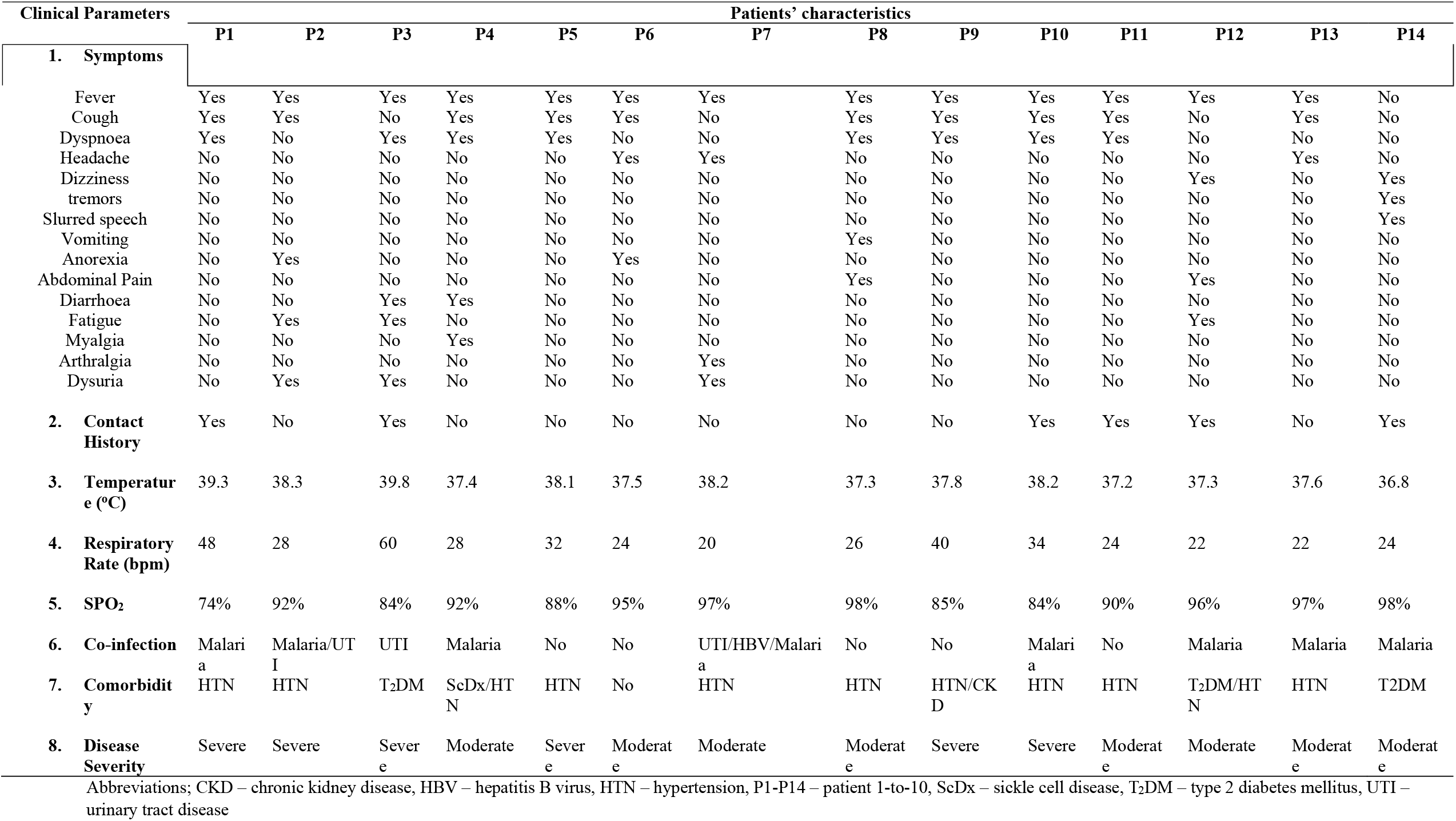
Clinical parameters and patient characteristics at presentation.

Following RT-PCR confirmation of cases, patients were classified as either having moderate or severe disease. Case classification was made based on the COVID-19 clinical classification of the Handbook of COVID-19 Prevention and Treatment [11]. Those with moderate cases were 8 (57.0%) and the remaining 6 (43.0%) had severe cases. During admission, two of the patients (one with moderate and the other with severe cases), progressed to critical disease (14%). Of the 14 cases reported, 13 (93%) had comorbidity (underlying non-communicable disease); eight (62%) had hypertension alone, one each had hypertension and type-2 diabetes mellitus (7.7%), hypertension, and sickle cell disease (7.7%) and hypertension and chronic kidney disease (7.7%), two (15%) had type-2 diabetes mellitus (T_2_DM) alone. Also, co-infection (e.g. malaria) was present in nine (64.3%) of the 14 cases as detailed in table 1. At presentation, all patients with T_2_DM, presented with hyperglycaemia. One of the patients who is not a known T_2_DM patient, presented with hyperglycaemia as shown in table 2. Likewise, seven (50%) out of the 14 cases, presented with leucocytosis; six of the seven had lymphopenia. Other details of the patients’ complete blood count are presented in table 2. Figure 1 also, shows the chest radiograph of the cases with imaging signs of pulmonary involvement.

**Table 2:**
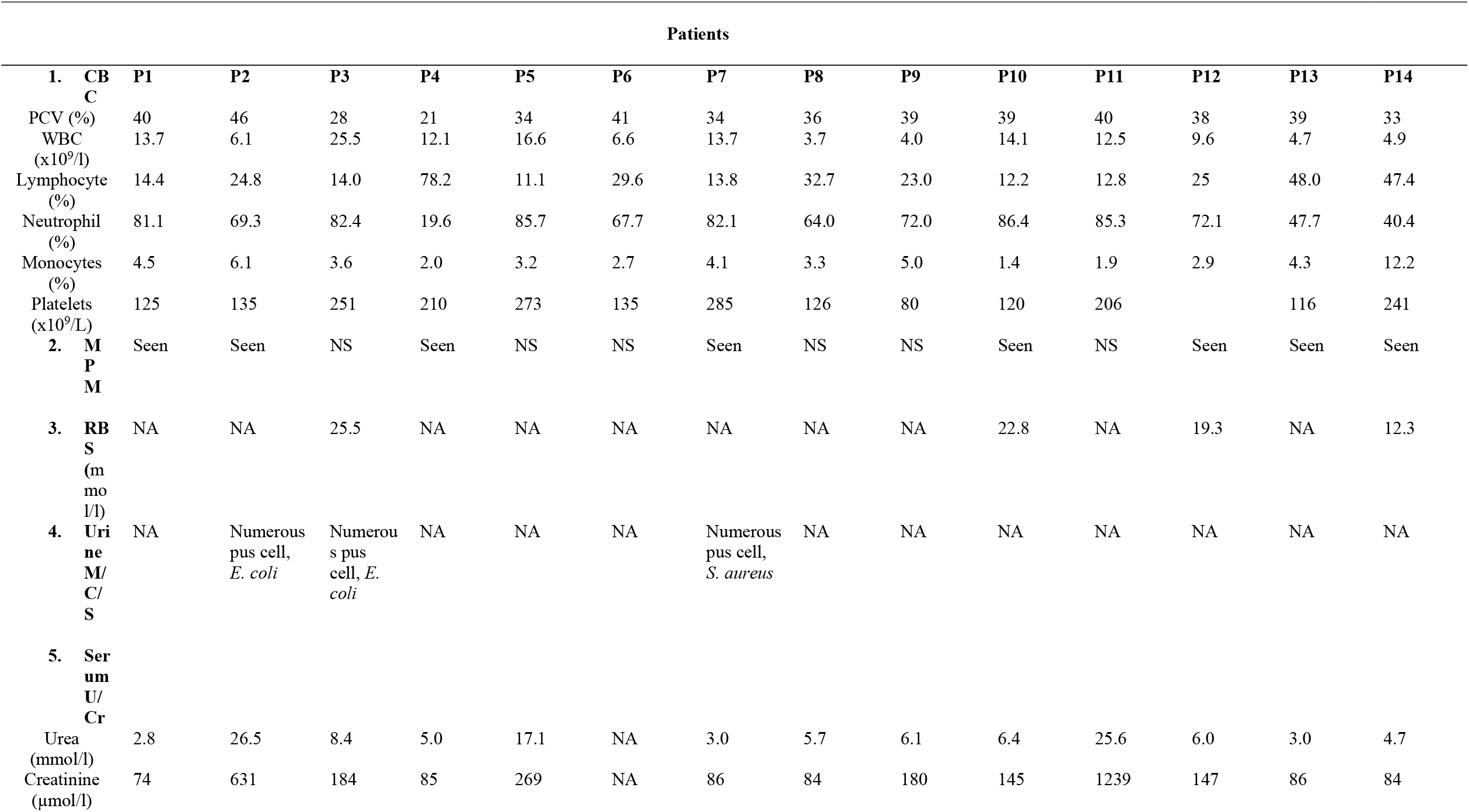
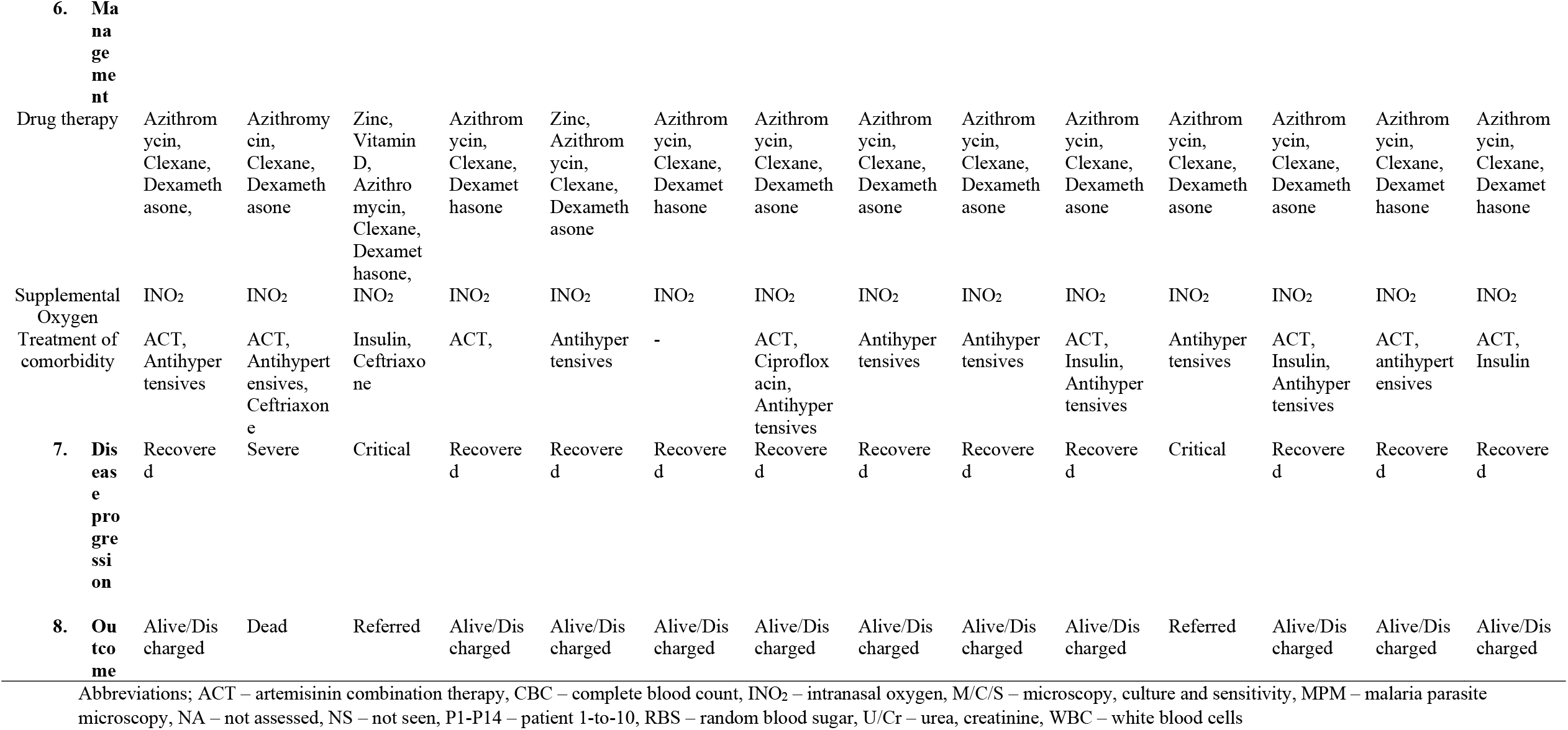
Patient evaluation, progression, Management, and outcome

**Figure 1:**
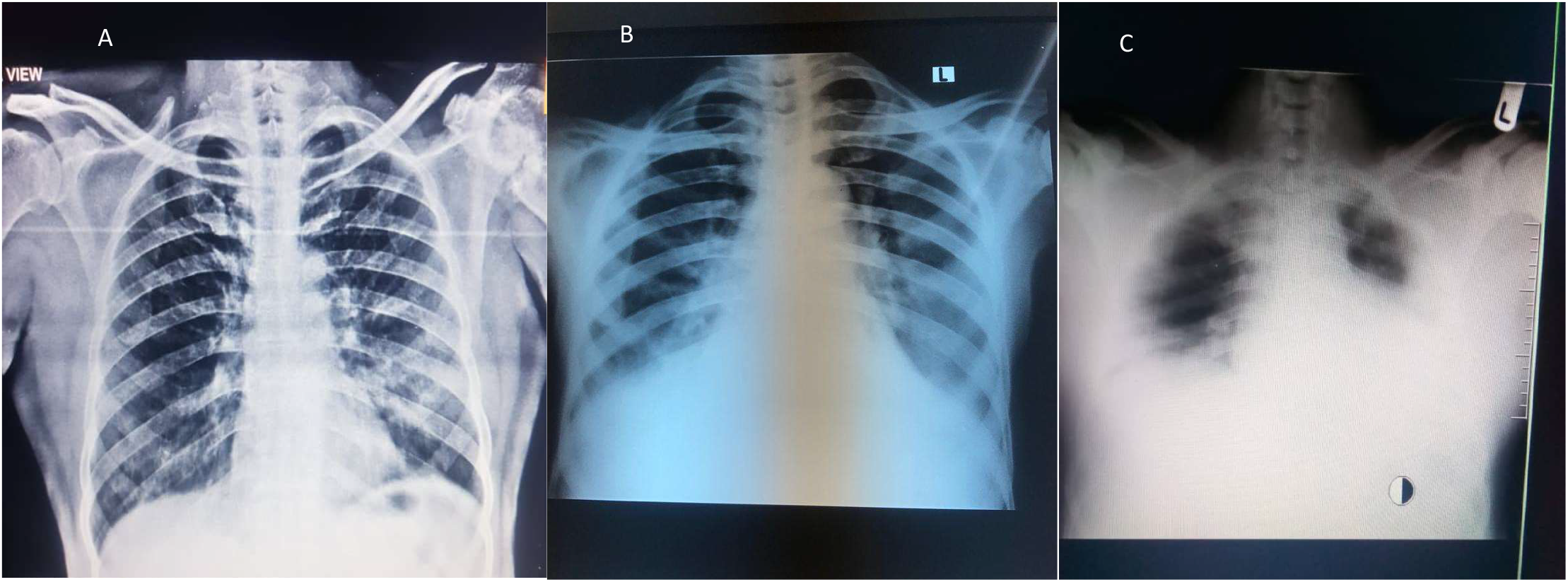
Chest radiograph of selected patients **A**; frontal chest radiograph of a 65 year old man with COVID-19 pneumonia showing bilateral peripheral opacity more prominent on the left side. **B;** frontal chest radiograph of a 60 year old male COVID-19 patient showing prominence of the central pulmonary vasculature with faint bilateral peripheral opacity more on the left side. **C;** a 43 year old male COVID-19 patient with bilateral lower lobe pneumonic consolidation. The image shows bilateral lower zone air space shadowing obscuring the diaphragmatic outline and the lower cardiac border.

All patients were treated with azithromycin, dexamethasone, and clexane (enoxaparin; a low-molecular-weight heparin). Some received additional treatment of zinc and vitamin D (Table 2 and 3). Patients were also given supplemental oxygen via an intranasal catheter. Comorbidities and co-infections were treated accordingly (Table 2). Two of the patients developed acute kidney injury complications which necessitated renal replacement therapy (haemodialysis). The two patients had progression of their clinical condition to critical disease thus, were referred for intensive care (ICU) management. Both patients died at the referral facilities (2 and 3 days after referral). Of the remaining 12 patients, one died and 11 (92%) recovered and were discharged alive from the hospital. The discharge criteria were clinically based on at least three days symptoms-free or sustained improved room air SPO2 of 95% for at least three days [10]. The median duration of hospital stay for the patients was 13 days (IQR; 8.25-15.0).

**Table 3:** Drug regimen for COVID-19 treatment

| S/N | Drugs | Route of administration | Dosage | Frequency | Duration |
| --- | --- | --- | --- | --- | --- |
| 1 | Azithromycin | Oral | 500mg/250mg | 500mg loading dose then 250mg daily | 3 – 5 days |
| 2 | Dexamethasone | Oral | 6mg | Once | 7 – 10 days |
| 3 | Claxane | Subcutaneous | 40mg (0.4ml) | Once | 5 – 14 days |
| 4 | Zinc | Oral | 100mg | Once | 10 days |
| 5 | Vitamin D | Oral | 4000IU | Once | 10 days |

## Discussion

There is limited information describing the clinical characteristics and outcome of admitted COVID-19 patients in Nigeria. Even though the availability of this information is essential to achieve effective control against COVID-19 in Nigeria and Africa. Thus, this case series will add to the few available reports on the much-needed COVID-19 clinical data in Nigeria. This case series reported 14 patients admitted and managed for COVID-19 at IBBSH in Nigeria. As reported in different studies [12] that the elderly are the group with a high prevalence of admission across the globe also, in this case series, all are in the age range of 42 – 80 years (mean age 63.5 [SD: 11.5]). This pattern of hospitalisation, conforms with reports from other parts of the world which shows that increased age is a major risk factor for hospitalisation in COVID-19 patients [13–15]. In consistence with previous studies, this report also shows that more males are likely to be hospitalised than females [16]. Where 78.6% of cases presented in this study are males. The reason for this gender disparity is still a subject of debate. Some researchers have attributed this to a higher concentration of the angiotensin converting enzyme 2 (ACE2) in testes tissue and plasma in males [17, 18]. Another explanation is that females tend to have a stronger immune response due to the protective effect of some genes expressed by chromosome X [19].

Also worthy of mention is the contact history of patients in this study. One of the patients that presented to the hospital during the early phase of the pandemic in Nigeria, had a history of travel. This goes to support the narration that most early cases of the infection in Nigeria were either imported cases or those in contact with the imported cases [20]. Another important dimension of the contact history is that four of the patients had a history of contact with hospitals where confirmed cases have been reported earlier. These facilities were not designated COVID-19 treatment centres. Perhaps many cases like these might have gone unnoticed and may have led to high cases of community transmission of the disease in the country. The public health significance of identifying this form of contact history is that it will help in mitigation and control of community spread through adequate contact tracing.

The presenting symptoms of the reported cases were also evaluated in this study. The commonest presenting symptoms were fever, cough, and dyspnoea. These symptoms have been reported in several studies across the globe as the commonest presenting symptoms [21]. Although in this study, fever is the most reported symptom seen in 13 out of the 14 patients. This is similar to what was reported in an earlier study in Lagos of the first 32 COVID-19 patients in Nigeria [22]. However, some studies have reported cough as the commonest presenting symptom [23, 24]. Nevertheless, the predominance of fever in this case series might not be unconnected with the occurrence of co-infection as observed in the cases. In all, nine cases (64.3%) presented with co-infection, and malaria (89%) was the most prevalent infection identified. In some of the cases, two co-infections (UTI and malaria) exist which will further support the fever predominance. This shows that clinical signs and symptoms might not be specific for COVID-19. The information has public health significance for policy planning and intervention to achieve effective control of the disease. Also, at the hospital level, the need for holistic patient evaluation by clinicians in identifying existing co-infections cannot be over-emphasised. Additionally, clinicians should also be on the lookout for less common and non – specific symptoms (Table 1). This will assist in instituting prompt and adequate treatment to reduce morbidity and mortality.

In this study, six out of the 14 patients presented with severe disease. While the remaining eight had a moderate disease. Similar to previous studies [25–28], all those with severe disease in this series were age 60 years and above. Already, advanced age has been shown to be a complex and multifactorial process [25]. Coexisting diseases are also common in the elderly which may predispose them to have severe COVID-’19. Similarly, in this report 13 of the 14 patients had underlying health conditions. Hypertension is the commonest comorbidity and this is not dissimilar with what has been reported in earlier studies in Nigeria as well as other parts of the world [26, 29, 30]. Two of the cases progressed to a critical disease requiring mechanical ventilation and ICU management. Background comorbidity might have played a role in the clinical course of the two cases. One had a pre-existing T_2_DM and the other had hypertension. The two patients thus developed acute (on chronic) kidney injury during management, and one was dialysed before referral. Their deterioration might not be unconnected with the development of the kidney disease complication. Studies have shown that kidney disease is associated with an increased risk of hospitalisation, critical COVID-19 infection, and fatal outcomes [31, 32]. Another prevalent co-existing disease condition in this report is T_2_DM. All the T_2_DM patients presented with hyperglycaemia (Table 2) despite being regular on their antidiabetic medications. Hyperglycaemia was also found in one of the patients who was previously not known to have T_2_DM. Indicating that the hyperglycaemic state might have been triggered by the SARS-CoV-2 infection. Several studies have suggested the bi-directional relationship between COVID-19 and T_2_DM (or new – onset diabetes) [33]. This has been attributed to many reasons which include the diabetogenic effect of COVID-19, stress response linked with severe illness, or glucocorticoid treatment [33].

It is also important to note that in this report, those with the severe disease tend to have more of leucocytosis, lymphopenia, neutrophilia, and thrombocytopenia (Table 2). While those with moderate infection have more of the normal haematologic findings. The haematologic picture seen here is in tandem with earlier published literature [34]. Likewise, both lymphopenia and neutrophilia have been associated with an increased risk of severe disease [35]. In some reports also, neutrophilia and lymphopenia were shown to be strong predictors of severe disease and ICU admission [36]. The mechanism leading to lymphopenia and neutrophilia in COVID-19 patients could be multidimensional. The direct cytopathic effect, increased apoptosis (for lymphopenia), and deranged immune homeostasis or secondary bacterial infections (for neutrophilia) have all been proposed as possible reasons [35]. This information might be of high significance for clinical monitoring and evaluation of COVID-19 patients. The haematological findings can serve as predictors for poor disease prognosis and as indicators for monitoring patient recovery [35]. Patients were also evaluated with chest radiography and the chest radiograph revealed findings that are consistent with COVID-19. The images in this report, revealed bilateral peripheral opacity in two of the patients while one had features of pneumonic consolidation (Figure 1A-C). These findings are suggestive of COVID-19 pneumonia as reported in previous studies [37, 38]. Although features of pneumonia on chest radiograph are not diagnostic of COVID-19, findings of such might be helpful in the diagnosis of symptomatic patients. Especially in resource-limited regions with challenges of diagnostic tools and where patients often present late.

Patients were treated with a combination of azithromycin, dexamethasone, and clexane or some, with the addition of zinc and vitamin D. The outcome in this report after treatment was impressive when compared to results of studies from some other parts of the world [26, 39]. Of the 14 patients, two (14%) had progression of disease severity thus, recuring ICU management and mechanical ventilation. The two patients were referred, and both died at the referral hospitals. However, of the remaining 12 patients one died (8%), and all others (92%) recovered and were discharged alive. The patient that died was an elderly hypertensive with a history of two previous stroke. These multiple factors have all been identified to be poor prognostic features in COVID-19 patients. Also, the 4 days median duration of symptoms onset to admission in this report was similar to reports from other studies [40]. Similarly, the median duration of hospital stay (13 days) of cases seen here is akin to what has been reported in other studies [22].

This case series has presented a comprehensive clinical detail of COVID-19 patients in Nigeria. The study has provided the much-needed COVID-19 clinical information from Nigeria. Thus, providing Nigeria and indeed Africa’s perspective of COVID-19 patients’ outcome towards the effective control of the pandemic. However, the study is not without limitations. The study is limited in the number of included patients. Since only one hospital was considered and only hospitalised patients with complete medical records were included. Additionally, information on mild and asymptomatic confirmed cases was not evaluated. Therefore, more multicentred studies that will evaluate the entire spectrum of the disease severity are required to address these challenges.

In conclusion, this study has provided evidence that as in other regions of the world, advanced age, male gender, and comorbidities are factors that increase the risk of hospitalisation in COVID-19 patients. Also, patients presenting symptoms, duration of symptoms onset to admission, and length of hospitalisation of Nigerian patients are like those reported from across the world. However, disease severity and survival outcome of patients from this study are better when compared to similar studies from Europe and some parts of the world.

## Data Availability

Data used to support the findings of this study were obtained from patients hospital records and are only available from the corresponding author on reasonable demand.

## Data Availability Statement

Data used to support the findings of this study were obtained from patient’s hospital records and are only available from the corresponding author on reasonable demand.

## Author Contributions

Conceptualization, B.W, S.H, & Y.R.; data extraction and analysis, Y.R, S.H, & A.J; writing—original draft preparation, Y.R.; writing—review and editing, B.W, S.H, A.J, U.A, A.A, A.M, & A.S; project administration, B.W, U.A.; funding acquisition, All authors contributed to the funding. All authors have read and agreed to the published version of the manuscript.

## Consent

No written consent has been obtained from the patients as there is no patient identifiable data included in this case series.

## References

1. Lu, H., C.W. Stratton, and Y.W. Tang, Outbreak of pneumonia of unknown etiology in Wuhan, China: the mystery and the miracle. Journal of medical virology, 2020. 92(4): p. 401–402.

2. Wang, H., et al., The genetic sequence, origin, and diagnosis of SARS-CoV-2. European Journal of Clinical Microbiology & Infectious Diseases, 2020. 39(9): p. 1629–1635.

3. WHO, W.H.O., Novel Coronavirus (2019-nCoV): situation report, 22. 2020.

4. Gorbalenya, A.E., et al., The species Severe acute respiratory syndrome-related coronavirus: classifying 2019-nCoV and naming it SARS-CoV-2. Nature Microbiology, 2020. 5(4): p. 536–544.

5. Zhu, N., et al., A Novel Coronavirus from Patients with Pneumonia in China, 2019. The New England journal of medicine, 2020. 382(8): p. 727–733.

6. Rothan, H.A. and S.N. Byrareddy, The epidemiology and pathogenesis of coronavirus disease (COVID-19) outbreak. Journal of Autoimmunity, 2020. 109: p. 102433.

7. NCDC, An update of COVID-19 outbreak in Nigeria, in COVID-19 OUTBREAK IN NIGERIA: Situation Report. 2020, Nigeria Center for Disease Control.

8. NCDC. COVID-19 Nigeria - Nigeria Centre for Disease Control. [cited 2021 20/03]; Available from: https://covid19.ncdc.gov.ng/.

9. World Health, O., Clinical management of COVID-19: interim guidance, 27 May 2020. 2020, World Health Organization: Geneva.

10. NCDC, National Interim Guidelines for Clinical Management of COVID-19, N.C.f.D. Control, Editor. 2020, Nigeria Centre for Disease Control.

11. Handbook of COVID-19 Prevention and Treatment, L. Tingbo, Editor. 2020, The First Affiliated Hospital, Zhejiang University School of Medicine: China. p. 68.

12. Richardson, S., et al., Presenting Characteristics, Comorbidities, and Outcomes Among 5700 Patients Hospitalized With COVID-19 in the New York City Area. JAMA, 2020. 323(20): p. 2052–2059.

13. Giorgi Rossi, P., et al., Characteristics and outcomes of a cohort of COVID-19 patients in the Province of Reggio Emilia, Italy. PLoS One, 2020. 15(8): p. e0238281.

14. Killerby, M.E., et al., Characteristics Associated with Hospitalization Among Patients with COVID-19 - Metropolitan Atlanta, Georgia, March-April 2020. MMWR. Morbidity and mortality weekly report, 2020. 69(25): p. 790–794.

15. Soares, R.d.C.s.M., L.R. Mattos, and L.c.M. Raposo, Risk Factors for Hospitalization and Mortality due to COVID-19 in Esp?rito Santo State, Brazil. The American Journal of Tropical Medicine and Hygiene, 2020. 103(3): p. 1184–1190.

16. Garg, S., et al., Hospitalization rates and characteristics of patients hospitalized with laboratory-confirmed coronavirus disease 2019—COVID-NET, 14 States, March 1–30, 2020. Morbidity and mortality weekly report, 2020. 69(15): p. 458.

17. Sama, I.E., et al., Circulating plasma concentrations of angiotensin-converting enzyme 2 in men and women with heart failure and effects of renin–angiotensin–aldosterone inhibitors. European heart journal, 2020. 41(19): p. 1810–1817.

18. Hamming, I., et al., Tissue distribution of ACE2 protein, the functional receptor for SARS coronavirus. A first step in understanding SARS pathogenesis. The Journal of Pathology: A Journal of the Pathological Society of Great Britain and Ireland, 2004. 203(2): p. 631–637.

19. Jaillon, S., K. Berthenet, and C. Garlanda, Sexual dimorphism in innate immunity. Clinical reviews in allergy & immunology, 2019. 56(3): p. 308–321.

20. Olapoju, O.M., Estimating transportation role in pandemic diffusion in Nigeria: A consideration of 1918-19 influenza and COVID-19 pandemics. Journal of Global Health, 2020. 10(2).

21. Chen, T., et al., Clinical characteristics of 113 deceased patients with coronavirus disease 2019: retrospective study. bmj, 2020. 368.

22. Bowale, A., et al., Clinical presentation, case management and outcomes for the first 32 COVID-19 patients in Nigeria. The Pan African Medical Journal, 2020. 35(24).

23. Abayomi, A., et al., Presenting Symptoms and Predictors of Poor Outcomes Among 2,184 Patients with COVID-19 in Lagos State, Nigeria. International journal of infectious diseases : IJID : official publication of the International Society for Infectious Diseases, 2021. 102: p. 226–232.

24. Macera, M., et al., Clinical presentation of COVID-19: case series and review of the literature. International Journal of Environmental Research and Public Health, 2020. 17(14): p. 5062.

25. Wei, C., et al., Clinical characteristics and manifestations in older patients with COVID-19. BMC Geriatrics, 2020. 20(1): p. 395.

26. Richardson, S., et al., Presenting characteristics, comorbidities, and outcomes among 5700 patients hospitalized with COVID-19 in the New York City area. Jama, 2020. 323(20): p. 2052–2059.

27. Abbatecola, A.M. and R. Antonelli-Incalzi, Editorial: COVID-19 Spiraling of Frailty in Older Italian Patients. The journal of nutrition, health & aging, 2020. 24(5): p. 453–455.

28. Huang, C., et al., Clinical features of patients infected with 2019 novel coronavirus in Wuhan, China. The Lancet, 2020. 395(10223): p. 497–506.

29. Otuonye, N.M., et al., Clinical and Demographic Characteristics of COVID-19 patients in Lagos, Nigeria: A Descriptive Study. Journal of the National Medical Association, 2020.

30. Zhou, F., et al., Clinical course and risk factors for mortality of adult inpatients with COVID-19 in Wuhan, China: a retrospective cohort study. The lancet, 2020. 395(10229): p. 1054–1062.

31. Henry, B.M. and G. Lippi, Chronic kidney disease is associated with severe coronavirus disease 2019 (COVID-19) infection. International urology and nephrology, 2020. 52(6): p. 1193–1194.

32. Kooman, J.P. and F.M. van der Sande, COVID-19 in ESRD and Acute Kidney Injury. Blood purification, 2020: p. 1–11.

33. Sathish, T., et al., Proportion of newly diagnosed diabetes in COVID-19 patients: a systematic review and meta-analysis. Diabetes, obesity & metabolism, 2020.

34. Selim, S., Leukocyte count in COVID-19: an important consideration. The Egyptian Journal of Bronchology, 2020. 14(1): p. 1–2.

35. Henry, B.M., et al., Lymphopenia and neutrophilia at admission predicts severity and mortality in patients with COVID-19: a meta-analysis. Acta Bio Medica: Atenei Parmensis, 2020. 91(3): p. e2020008.

36. Lippi, G. and M. Plebani, Laboratory abnormalities in patients with COVID-2019 infection. Clinical Chemistry and Laboratory Medicine (CCLM), 2020. 58(7): p. 1131–1134.

37. Cleverley, J., J. Piper, and M.M. Jones, The role of chest radiography in confirming covid-19 pneumonia. bmj, 2020. 370.

38. Brennan, Z., S. Guerra, and S. Seman, Radiological Findings of COVID-19 Patients in Italy. Spartan Medical Research Journal, 2020. 5(2).

39. Guglielmetti, L., et al., Severe COVID-19 pneumonia in Piacenza, Italy — A cohort study of the first pandemic wave. Journal of Infection and Public Health, 2021. 14(2): p. 263–270.

40. Docherty, A.B., et al., Features of 20 133 UK patients in hospital with covid-19 using the ISARIC WHO Clinical Characterisation Protocol: prospective observational cohort study. bmj, 2020. 369.

